# Raising the patient’s voice in influenza research and care with patient-reported outcomes

**DOI:** 10.1101/2025.04.15.25325874

**Authors:** Elizabeth Meda-Monzón, Paloma Muñoz-Aguirre, Barbara Aída Flores-Aldana, Chad Gwaltney, José Bartelt-Hofer

**Affiliations:** Pharmamanagement, Mexico City, Mexico; Population Health Research Center, National Institute of Public Health, Cuernavaca, Mexico; Gwaltney Consulting, Westerly, RI, USA; Sanofi, Lyon, France

**Author notes:** **Correspondence:** José Bartelt-Hofer, Senior Project Lead, Global Health Economics and Outcomes Research, Sanofi, 14 Espace Henry Vallée, Lyon 6900712, France.

**Keywords:** Influenza, influenza-like illness, influenza interventions, patient-reported outcomes, patient-reported outcomes instruments, patient care, routine care, clinical research, guidelines

## Abstract

**Background:** Patient-reported outcomes (PROs) enable the evaluation of patients’ perspectives in relation to their health and well-being.

**Methods:** We conducted a systematic literature review of publications and electronic records reporting PROs for influenza and influenza-like illness.

**Results:** The systematic search identified 112 items, of which 36 were included in the final analysis (20 clinical research publications; two national guidelines; eight influenza intervention product labels; and six PRO instrument descriptions). We identified influenza-specific PRO instruments developed to assess symptom severity and impact of influenza, or the acceptability and safety of influenza interventions; several of these have been used in clinical research, enabling a better understanding of the impact of influenza and influenza interventions. However, further research on the use of influenza-specific PROs in routine care settings is needed.

**Conclusions:** Based on these findings, we describe opportunities and challenges for PROs and propose recommendations to facilitate wider implementation of PROs in influenza research and patient care.

## Introduction

Influenza is an acute respiratory infectious disease that spreads easily, with rapid transmission in crowded areas (1, 2). Influenza affects hundreds of millions of individuals worldwide causing substantial morbidity, with an estimated 294,000 to 518,000 global deaths caused by influenza each year, equating to approximately 2% of all annual respiratory deaths (3). Acute influenza infection can result not only in a high symptom burden for individuals but can also cause a wide range of severe complications including exacerbation of underlying chronic conditions and increased risk of diseases across organ systems, such as cardiovascular disease (4, 5).

Patient-reported outcomes (PROs) are a category of health-related endpoints that rely on direct input from patients and can encompass various aspects of their lived experience of conditions, including health status and overall well-being, symptoms, functional abilities, emotional state, quality of life (QoL), and treatment satisfaction (6, 7). PROs are of fundamental importance to healthcare, as they serve as a valuable means of capturing a patient’s perspective on their health and healthcare experiences, thereby enabling healthcare professionals to provide a more personalized and holistic approach to care and allowing health authorities to make more informed public health decisions (6). Furthermore, guidance documents from health authorities inform the development, selection, and use of PROs in clinical trials as a way of fostering patient-centered drug and vaccine development (6, 8).

There are numerous examples of generic health status measures that are used across clinical research and by health authorities, such as the widely used EuroQol five dimensions (EQ-5D) and the 36-item short form health survey (SF-36) in their multiple forms (9–11). Despite the value of such generic PRO instruments, which can allow for the comparison of endpoints across multiple heterogenous indications, they are designed to assess general aspects of health, and so are less sensitive to the capture of disease-specific attributes, potentially underestimating the real value of interventions (7, 12). Several disease-specific PRO measures are available for the evaluation of some conditions, for example, the European Organization for Research and Treatment of Cancer QLQ-30 (13), and the Hemophilia Treatment Experience Measure (Hemo-TEM) (14). Use of more targeted PRO instruments such as these may allow for more detailed evaluation of the true impact on patients of a specific disease or condition (15). Adding a patient-centered perspective to bridge understanding with traditional measures for influenza is particularly important given that patient experiences of influenza can vary substantially from person-to-person (1).

This paper provides a comprehensive review of PROs and PRO instruments used in the context of influenza or influenza-like illness (ILI). Our systematic approach allowed for consideration of the topic from a wide perspective, which included a review of the available PRO instruments, examples of their use in clinical research and by health authorities, and potential further opportunities for research.

## Methods

### Eligibility criteria

In this systematic literature review, research studies, electronic records, or regulatory documents that have mentioned or provided descriptions of the use of PRO instruments in the context of influenza or ILI were eligible for inclusion.

### Information sources and search strategy

A systematic search was conducted from June to October 2023 across the MEDLINE database and COA-specific databases from the ePROVIDE platform (PROQOLID, PROLABEL, and PROINSIGHT). The databases within the ePROVIDE platform enabled collection of data on PROs in influenza research studies, and regulatory guidelines for their utilization and inclusion in product labels for influenza interventions. Following the PICO (Population, Intervention, Comparison, Outcome) framework,(55) the following Medical Subject Heading (MeSH) terms and keywords were chosen for the search algorithms: “influenza”, “influenza, human”, “influenza vaccines”, “patient-reported outcome measures”, “influenza-like illness”, “Influenza Impact Wellbeing Scale”, “Influenza Symptom Severity Scale”, “vaccinees’ perception of injection”, “Influenza Intensity and Impact Questionnaire”, “Respiratory Intensity and Impact Questionnaire”, and “InFLUenza Patient-Reported Outcome”.

### Study selection and data extraction

Two reviewers (EMM and PMA) conducted independent assessments of all references according to the search eligibility criteria. Full texts of references were comprehensively screened to identify relevant literature. Any discrepancies or disputes regarding eligibility were resolved through discussion between the two reviewers. The information extracted from the identified articles included: publication year, objective(s), target population, instrument, covered domains, timepoint collection, and principal findings.

## Results

### Study selection and characteristics

The literature search identified a total of 112 items: 57 articles from MEDLINE and 55 items (17 articles; 38 electronic records) from the ePROVIDE platform (PROQOLID, PROLABEL, and PROINSIGHT). A comprehensive overview of the search process is shown in the PRISMA diagram (Fig. 1).

**Fig. 1.**
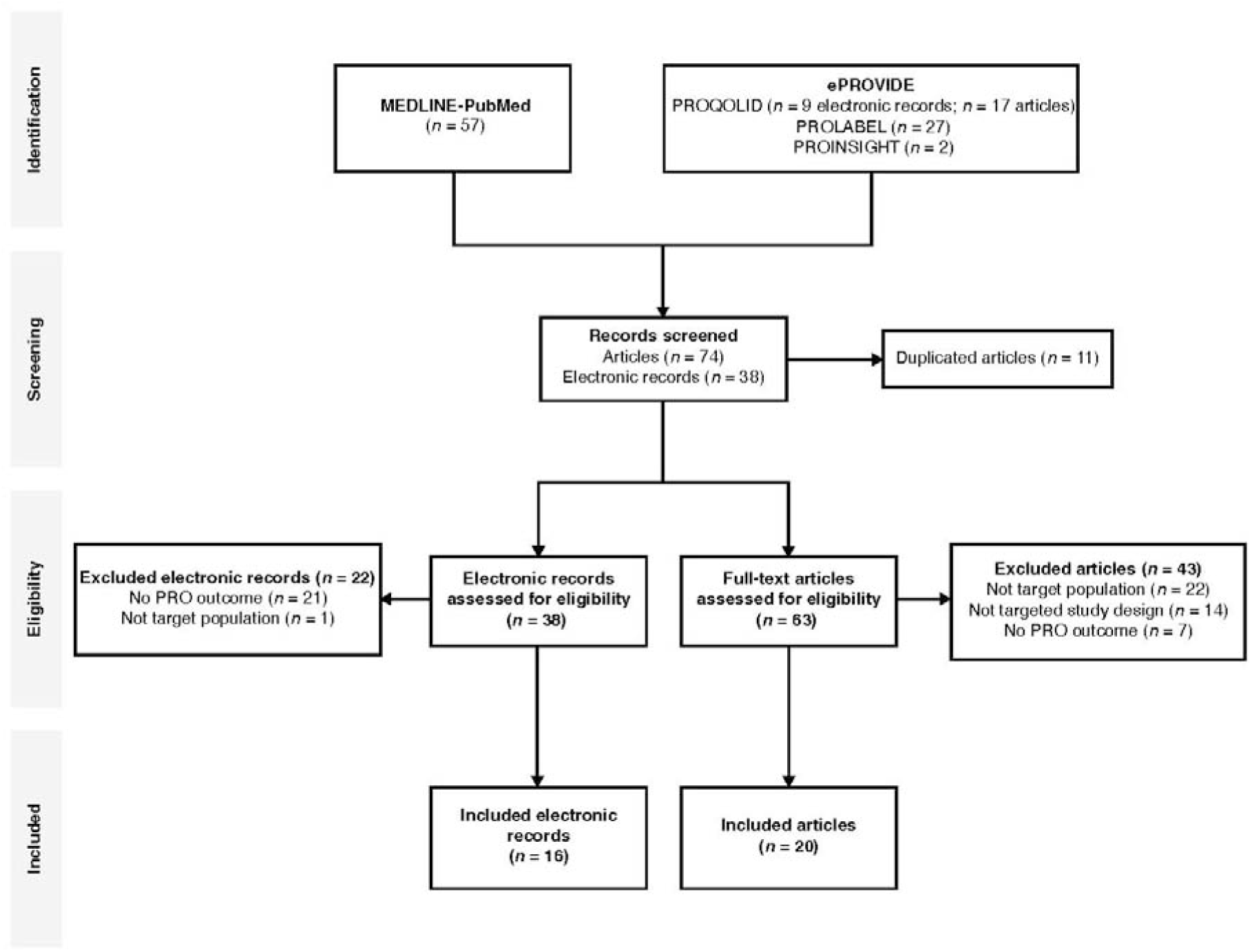
PRISMA flow diagram of the article selection process in the systematic literature review. PRO: patient-reported outcome

Following removal of duplicated articles (*n* = 11), application of the pre-specified eligibility criteria and a comprehensive full-text evaluation, 20 articles and 16 electronic records were included in the final analysis. Sixty-five records were excluded; the reasons for exclusion were: no PRO endpoint was included, the study design was not the target, and the population was not the target.

Two influenza-related instruments were excluded: the first because it measures observer-reported outcomes rather than PROs (the Canadian Acute Respiratory Illness and Flu Scale [CARIFS], developed to assess symptom severity and impact of acute respiratory diseases in children ≤12 years old and subsequent impact on their caregivers); the second because the caregiver was the target population rather than the patient (the Care-ILI-QoL questionnaire, developed to assess the quality of life of caregivers of children with ILI).

Of the 36 items included in the final analysis, 20 articles described the development, psychometric properties, or clinical application of PRO instruments (Table 1) (16–35), two were regulatory documents guiding the use of PROs in influenza clinical trial design (36, 37), and eight indicated the inclusion of PROs in the clinical section of product label claims (Table 2) (38–45). The remaining six records detailed digital descriptions of the eligible PROs; these records were used to detail the domain(s) of PRO instruments but are not discussed

**Table 1.**
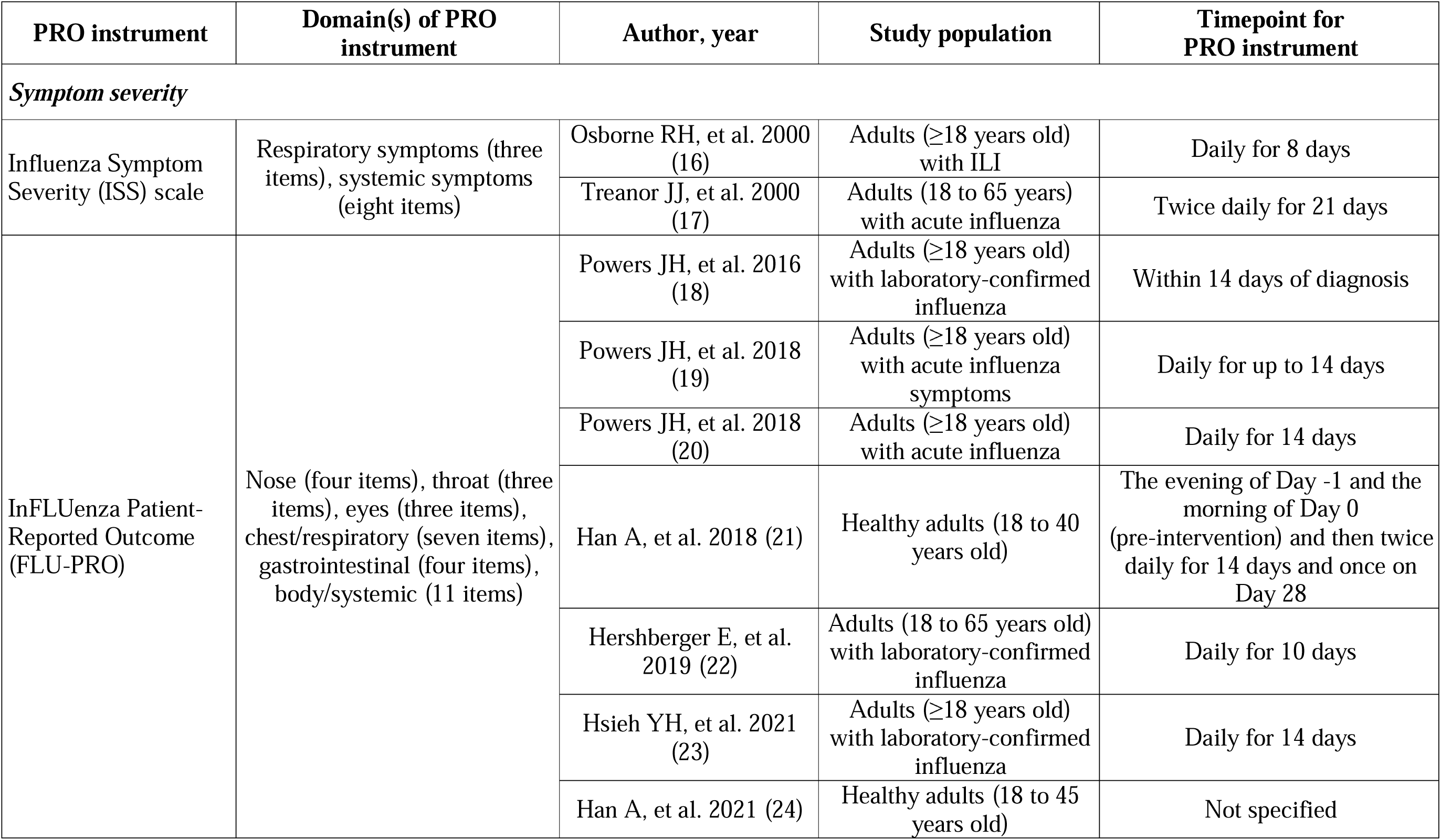
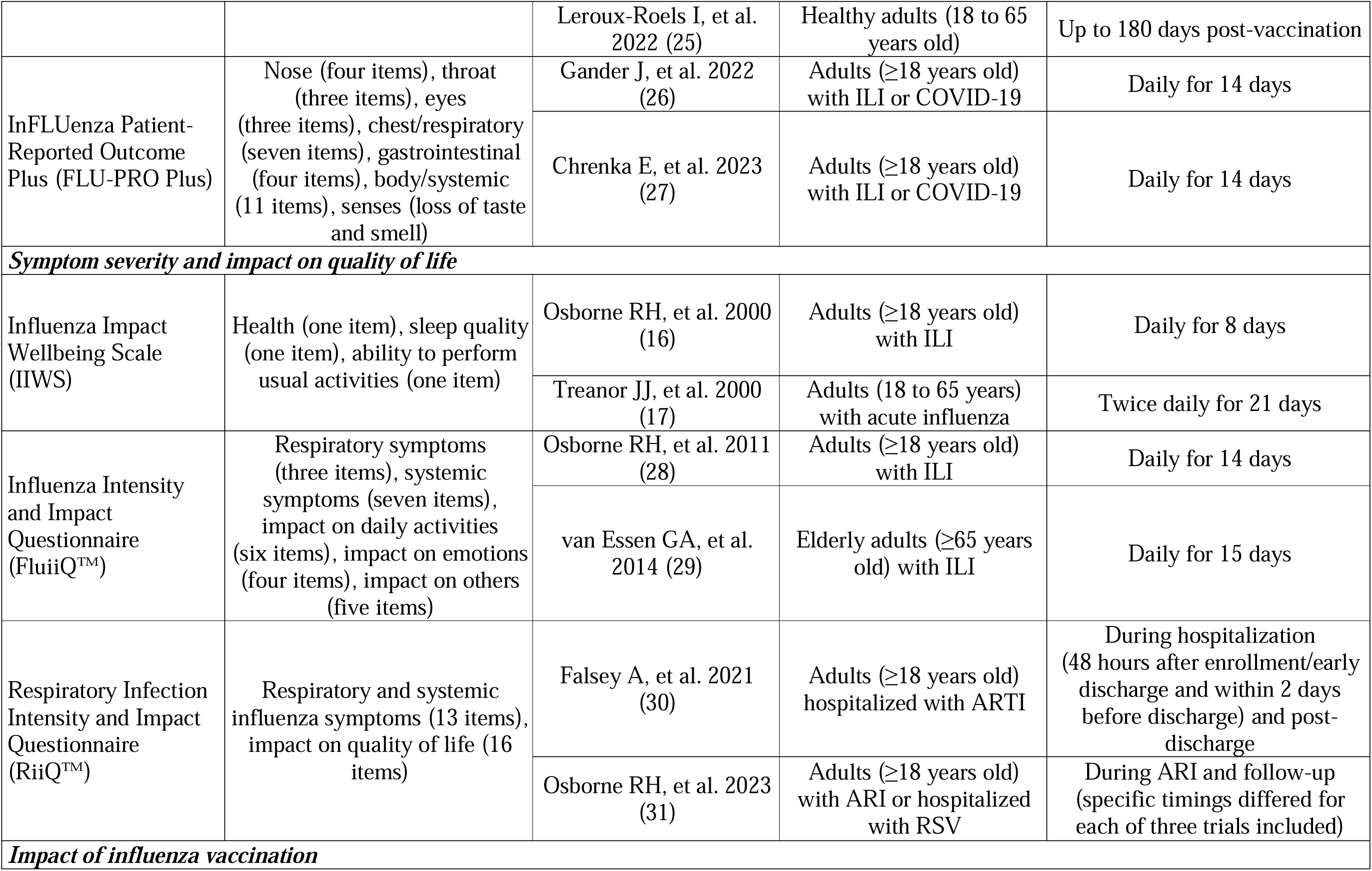
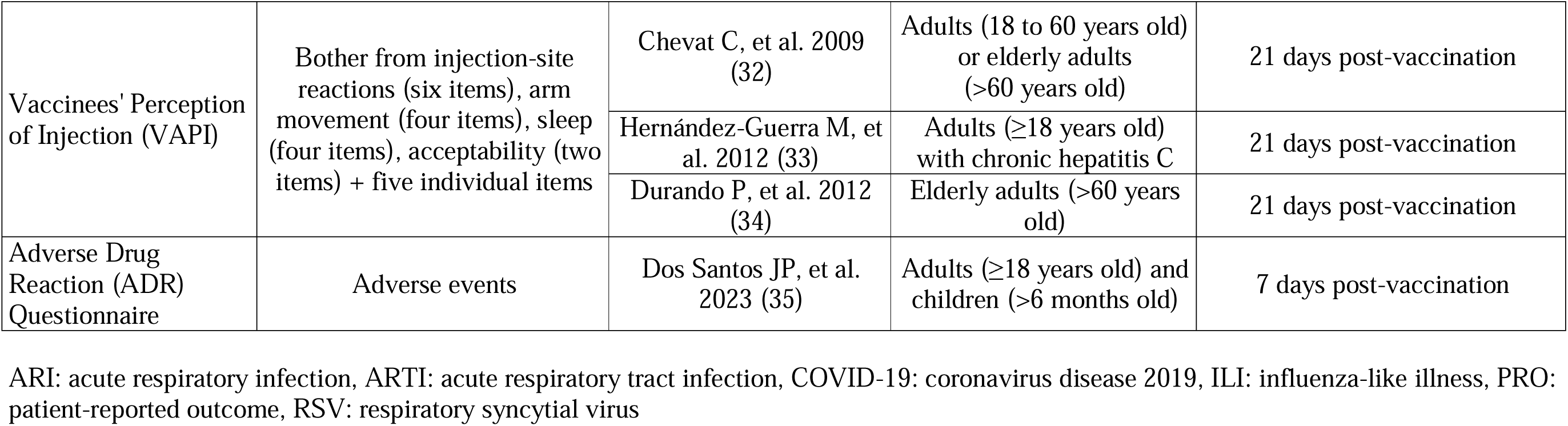
Identified PRO instruments used in influenza clinical research.

| PRO instrument | Domain(s) of PRO instrument | Author, year | Study population | Timepoint for PRO instrument |
| --- | --- | --- | --- | --- |
| *Symptom severity* |  |  |  |  |
| Influenza Symptom Severity (ISS) scale | Respiratory symptoms (three items), systemic symptoms (eight items) | Osborne RH, et al. 2000 (16) | Adults ($\geq 18$ years old) with ILI | Daily for 8 days |
|  |  | Treanor JJ, et al. 2000 (17) | Adults (18 to 65 years) with acute influenza | Twice daily for 21 days |
| InFLUenza Patient-Reported Outcome (FLU-PRO) | Nose (four items), throat (three items), eyes (three items), chest/respiratory (seven items), gastrointestinal (four items), body/systemic (11 items) | Powers JH, et al. 2016 (18) | Adults ($\geq 18$ years old) with laboratory-confirmed influenza | Within 14 days of diagnosis |
| | | Powers JH, et al. 2018 (19) | Adults ($\geq 18$ years old) with acute influenza symptoms | Daily for up to 14 days |
| | | Powers JH, et al. 2018 (20) | Adults ($\geq 18$ years old) with acute influenza | Daily for 14 days |
|  |  | Han A, et al. 2018 (21) | Healthy adults (18 to 40 years old) | The evening of Day -1 and the morning of Day 0 (pre-intervention) and then twice daily for 14 days and once on Day 28 |
|  |  | Hershberger E, et al. 2019 (22) | Adults (18 to 65 years old) with laboratory-confirmed influenza | Daily for 10 days |
| | | Hsieh YH, et al. 2021 (23) | Adults ($\geq 18$ years old) with laboratory-confirmed influenza | Daily for 14 days |
|  |  | Han A, et al. 2021 (24) | Healthy adults (18 to 45 years old) | Not specified |
|  |  | Leroux-Roels I, et al. 2022 (25) | Healthy adults (18 to 65 years old) | Up to 180 days post-vaccination |
| InFLUenza Patient-Reported Outcome Plus (FLU-PRO Plus) | Nose (four items), throat (three items), eyes (three items), chest/respiratory (seven items), gastrointestinal (four items), body/systemic (11 items), senses (loss of taste and smell) | Gander J, et al. 2022 (26) | Adults (≥18 years old) with ILI or COVID-19 | Daily for 14 days |
|  |  | Chrenka E, et al. 2023 (27) | Adults (≥18 years old) with ILI or COVID-19 | Daily for 14 days |
| Symptom severity and impact on quality of life |  |  |  |  |
| Influenza Impact Wellbeing Scale (IIWS) | Health (one item), sleep quality (one item), ability to perform usual activities (one item) | Osborne RH, et al. 2000 (16) | Adults (≥18 years old) with ILI | Daily for 8 days |
|  |  | Treanor JJ, et al. 2000 (17) | Adults (18 to 65 years) with acute influenza | Twice daily for 21 days |
| Influenza Intensity and Impact Questionnaire (FluIIQ™) | Respiratory symptoms (three items), systemic symptoms (seven items), impact on daily activities (six items), impact on emotions (four items), impact on others (five items) | Osborne RH, et al. 2011 (28) | Adults (≥18 years old) with ILI | Daily for 14 days |
|  |  | van Essen GA, et al. 2014 (29) | Elderly adults (≥65 years old) with ILI | Daily for 15 days |
| Respiratory Infection Intensity and Impact Questionnaire (RiiQ™) | Respiratory and systemic influenza symptoms (13 items), impact on quality of life (16 items) | Falsey A, et al. 2021 (30) | Adults (≥18 years old) hospitalized with ARTI | During hospitalization (48 hours after enrollment/early discharge and within 2 days before discharge) and post-discharge |
|  |  | Osborne RH, et al. 2023 (31) | Adults (≥18 years old) with ARI or hospitalized with RSV | During ARI and follow-up (specific timings differed for each of three trials included) |
| Impact of influenza vaccination |  |  |  |  |
| Vaccinees' Perception of Injection (VAPI) | Bother from injection-site reactions (six items), arm movement (four items), sleep (four items), acceptability (two items) + five individual items | Chevat C, et al. 2009 (32) | Adults (18 to 60 years old) or elderly adults (>60 years old) | 21 days post-vaccination |
| | | Hernández-Guerra M, et al. 2012 (33) | Adults ($\geq 18$ years old) with chronic hepatitis C | 21 days post-vaccination |
|  |  | Durando P, et al. 2012 (34) | Elderly adults (>60 years old) | 21 days post-vaccination |
| Adverse Drug Reaction (ADR) Questionnaire | Adverse events | Dos Santos JP, et al. 2023 (35) | Adults ($\geq 18$ years old) and children (>6 months old) | 7 days post-vaccination |

ARI: acute respiratory infection, ARTI: acute respiratory tract infection, COVID-19: coronavirus disease 2019, ILI: influenza-like illness, PRO: patient-reported outcome, RSV: respiratory syncytial virus

**Table 2.** Identified PRO instruments described in product labels of influenza interventions.

| **Regulatory agency** | **Drug** | **Approval year** | **PRO endpoint of interest** | **PRO instrument(s) utilized** |
| --- | --- | --- | --- | --- |
| US FDA | Relenza (zanamivir)<br>(38) | 1999 | Time to alleviation or improvement of influenza symptoms | Unknown PRO instrument (five symptoms) |
|  | Tamiflu (oseltamivir)<br>(39) | 1999 | Time to alleviation of influenza symptoms | Unknown PRO instrument (seven symptoms assessed using a 4-point scale, twice daily)<br>Unknown PRO instrument and assessment type |
|  | Rapivab (peramivir)<br>(40) | 2014 | Time to alleviation or resolution of influenza symptoms | Unknown PRO instrument (seven symptoms assessed using a 4-point scale, twice daily)<br>Unknown PRO instrument and assessment type |
|  | Xofluza (baloxavir marboxil) (41) | 2018 | Time to alleviation or improvement of influenza signs and symptoms | Unknown PRO instrument (seven symptoms assessed using a 4-point scale, twice daily) |
| EMA | Tamiflu (oseltamivir)<br>(42) | 2002 | Time to alleviation of influenza symptoms | Unknown PRO instrument and assessment type |
|  | Alpivab (peramivir)<br>(43) | 2018 | Time to alleviation or resolution of influenza symptoms | Unknown PRO instrument (seven symptoms assessed using a 4-point scale, twice daily)<br>Unknown PRO instrument and assessment type |
|  | Dectova (zanamivir)<br>(44) | 2019 | Fever, upper respiratory tract illness and total symptoms scores | Unknown PRO instrument and assessment type |
|  | Xofluza (baloxavir marboxil) (45) | 2022 | Time to alleviation or improvement of influenza symptoms | Unknown PRO instrument (seven symptoms)<br>Unknown PRO instrument (seven symptoms) |

EMA: European Medicine Agency, PRO: patient-reported outcome, US FDA: United States Food and Drug Administration

### PRO instruments in influenza clinical research

Several PRO instruments have been developed, undergone psychometric testing, and subsequently been used in influenza clinical trials (Table 1). These broadly fall into the following categories of assessment: influenza symptom severity (16–27), influenza symptom severity and the impact of influenza (16, 17, 28–31), and the impact of influenza interventions (32–35).

#### Assessing the severity of influenza

Influenza symptom severity scales are disease-specific PRO instruments that capture the intensity, frequency, and duration of symptoms associated with influenza (e.g. cough, nasal congestion, sore throat, headache, fever, muscle aches, and fatigue). Such tools for use in adults include the discontinued influenza symptom severity (ISS) scale (16, 17), and the more recently developed InFLUenza Patient-Reported Outcome (FLU-PRO) and FLU-PRO Plus questionnaires (18–20, 26). FLU-PRO is a six-domain (eyes, nose, throat, chest/respiratory, gastrointestinal, and body/systemic), 32-item questionnaire that was developed to examine influenza symptoms, and includes a 5-point Likert-type scale scoring system ranging from 0 to 4, to allow measurement of both symptom presence and severity (20). The FLU-PRO questionnaire has been used in multiple phase 2 studies, including a double-blind, placebo-controlled study on the safety and efficacy of a monoclonal antibody (VIS410) (22), and a randomized, double-blind study on the immunogenicity and safety of a nucleoprotein-based influenza vaccine (OVX836) (25). Results from clinical studies suggest that FLU-PRO scores are reliable and responsive to change, and FLU-PRO has high adherence and low responder burden (19–21). The FLU-PRO questionnaire was subsequently modified into the FLU-PRO Plus by adding a seventh domain for sense items (e.g. loss of sense of smell) during the COVID-19 pandemic (26). Using this modified questionnaire, three symptom clusters successfully discriminated patients with COVID-19 from non-COVID illness and were associated with quality of life and predicted symptom duration (27).

#### Assessing the severity and impact of influenza

Symptom severity scales have been developed to allow self-reporting of the impact of influenza on patients’ daily lives and emotions. These PRO tools include the Influenza Intensity and Impact Questionnaire (FluiiQ™), developed from the discontinued ISS scale and influenza impact wellbeing scale (IIWS) following thorough psychometric work, and the Respiratory Infection Intensity and Impact Questionnaire (RiiQ™), further evaluated and adapted from the FluiiQ™ (28, 30). The FluiiQ^TM^ and RiiQ^TM^ items are scored using a 4-point scale ranging from 0 to 3, with higher scores denoting worse symptoms and impact (28, 30).

The FluiiQ™ is a five-domain, 25-item PRO tool, which was developed through concept mapping with patients who had recently experienced influenza and through interviews with clinicians (28). The grounded concept mapping process allowed patients to report their full range of experience of influenza infection, and resulted in the generation of two symptom severity scales (respiratory and systemic) and three impact of influenza scales (impact on daily activities, emotions, and others) (28). The FluiiQ™ was found to have strong psychometric properties, including excellent evidence of construct validity, reliability, and responsiveness (28). It has been applied in a range of settings, including in randomized clinical trials to assess the efficacy of inactivated trivalent seasonal influenza vaccines (29).

As mentioned, the FluiiQ™ was subsequently further evaluated and adapted into the RiiQ™ to strengthen its application in a broader range of respiratory infections in adults (including respiratory syncytial virus) (30). The RiiQ™ consequently includes all scales used in FluiiQ^TM^ (symptom severity and impact of respiratory infection), but with a respiratory symptoms scale that has increased reliability (30). Using data from several clinical studies, the RiiQ™ has demonstrated construct validity, reliability, and responsiveness, including expected associations with clinical ratings and the EQ-5D-5L (30).

#### Assessing the safety and acceptability of influenza interventions

PRO tools have also been developed to assess the safety, perceptions, and acceptance of influenza interventions. This review identified the Vaccinees’ Perception of Injection (VAPI) questionnaire, developed to capture the most immediate side effects of vaccination and a patient’s general attitude to acceptability of vaccination (32–34), and an unknown adverse drug reaction (ADR) questionnaire, used once in the context of safety surveillance of an influenza vaccine (35).

The VAPI questionnaire is a self-administered questionnaire used to assess the acceptance of influenza vaccination and consists of domains measuring bother from injection-site reactions, arm movement, sleep, and acceptability (32). The psychometric properties of the VAPI questionnaire include face and construct validity, with its scales demonstrating good reliability for assessing the acceptability of vaccine injection (32). The VAPI questionnaire has subsequently been used in studies assessing the acceptability of influenza vaccines (32–34), although further confirmatory factor analysis would be beneficial to confirm the scoring algorithm and determine if items could be removed, prior to recommending its use.

The ADR questionnaire was a general questionnaire consisting of a comprehensive list of adverse events (35). Insufficient response scales, limited to the presence or absence of the event, and lack of details regarding its development and psychometric properties suggest that future enhancements are needed to the ADR questionnaire before its use in future clinical settings.

Additional information on the general purpose and procedures for validating and testing the reliability of PRO instruments, along with a description of the key psychometric properties of the PRO instruments described earlier from the papers in which the measures were originally developed, can be found in the Supplementary File S1 (18, 20, 28, 32).

### PRO instruments described in product labels and regulatory guidelines

This systematic review extended beyond clinical research studies to also identify descriptions of PROs in regulatory documents, specifically product labels for influenza interventions, limited to those from the United States Food and Drug Administration (FDA) and European Medicines Agency (EMA) (38–45), and regulatory guidelines for clinical trial design (36, 37).

As displayed in Table 2, the US FDA and EMA have each approved four influenza interventions over the period from 1999 to 2022 that reference PROs in the clinical section of the product labels. All identified results relate to interventions with the aim of relieving the symptoms of influenza infection (38–45). The most frequently reported PRO endpoint of interest among these product labels was time to improvement or alleviation of influenza symptoms, including cough, sore throat, nasal congestion, headache, feverishness, myalgia, and fatigue (38–41, 43, 45). In most studies described within the product labels, the self-reported PRO instrument was not identified, but often comprised tools that used a 4-point response scale range (e.g. “none”, “mild”, “moderate”, “severe”) (39–41, 43). Furthermore, frequency of completion of the PRO instrument by patients was also not commonly described, with only twice daily use of PRO instruments reported (39–41, 43), as opposed to once daily, once weekly, or any other possible frequencies used by other PRO instruments (20, 28).

The systematic literature search also identified two regulatory documents from the US FDA that described use of PROs in the design of influenza clinical trials(36, 37). In the US FDA (2011) regulatory document ‘*Influenza: Developing Drugs for Treatment and/or Prophylaxis*’, PROs are recommended for measuring the signs and/or symptoms of influenza as both primary (antiviral medications) and secondary (prophylaxis treatment) endpoints (36). In particular, industry sponsors are encouraged to provide evidence for the ability of interventions to directly measure how a patient feels, functions, or survives, potentially using self-assessments (e.g. use of diary cards) (36). In the US FDA (2021) regulatory document ‘*Clinical Outcome Assessment (COA) Compendium*’, PROs are recommended for consideration in trials assessing antivirals for the treatment of influenza (37).

## Discussion

In this systematic literature review, we utilized both a general database (MEDLINE) and COA-specific databases (PROQOLID, PROLABEL, and PROINSIGHT) to identify research studies or regulatory documents that mentioned or provided descriptions of the use of PRO instruments in the context of influenza or ILI. We identified articles and electronic records describing the development, validity testing, and use of PRO instruments in influenza clinical research and patient care, regulatory documents describing use of PROs in the design of influenza clinical trials, and several product labels for influenza interventions which mentioned the use of such instruments. Based on these findings, we can describe the advantages and challenges of using influenza-specific PRO instruments and provide recommendations for their wider implementation in influenza clinical research and care.

There are several advantages of using PROs in influenza clinical research and care. PROs allow/provide: 1) the early detection of complications and tracking of influenza (for example, by identifying signs of improvement or deterioration in individual patients), 2) a holistic understanding of the impact of influenza (including an assessment of what matters most to patients with influenza, and how they are feeling), 3) research insights for personalized patient care, and 4) endpoints that can be used in the development or safety monitoring of improved interventions for influenza.

Influenza symptoms often manifest suddenly, transiently, and can vary substantially from person to person (1, 2). The symptom severity PRO instruments (e.g. FLU-PRO) enable severity scoring of symptoms beyond just their presence and allow for continued active reporting of symptom severity from onset (e.g. once or twice daily) (18). Importantly, such instruments may enable the early detection of influenza complications or worsening of symptoms, allowing healthcare providers to promptly identify signs of deterioration and intervene in a timely manner to prevent progression of illness to a more severe stage.

PRO instruments can allow a holistic understanding of the impact of influenza on patients and provide research insights for personalized, patient-centered care, allowing healthcare professionals and researchers to understand the broader implications of influenza disease beyond physical signs and laboratory values (6). The development of influenza disease-specific tools such as the RiiQ™, which include measures for both symptom severity and the impact of influenza on the patient, are important for improving understanding of a patient’s condition and the support that they may require (30). Traditional trial metrics, such as infection rates, severe clinical outcomes (for example, hospitalization) or death, fail to capture the full impact of the disease on patients (7). Moreover, current safety monitoring often focuses solely on the presence or absence of adverse events, whereas PRO metrics provide a deeper understanding of the severity of symptomatic adverse reactions by using ordinal or continuous response scales (7). As a result, healthcare decisions may potentially become more collaborative between patients and healthcare professionals, and tailored to an individual’s needs (6, 46). Another feature of symptom severity PRO tools is that they allow comparison of symptoms of influenza with other respiratory diseases. For example, the RiiQ™ has been used to compare symptoms of influenza, respiratory syncytial virus, and human metapneumovirus (30) and the FLU-PRO Plus questionnaire was recently used to distinguish between COVID-19 and non-COVID-19 ILI in adults (26, 27). In doing so, these PRO tools have the potential to be implemented in joint respiratory disease trials or monitoring settings, facilitate comparisons of outcomes across respiratory diseases for decision-making, provide a supportive, preliminary identification of the disease in the absence of formal laboratory testing, and promote patient-centric, tailored treatments and interventions to alleviate the burden of specific respiratory diseases.

Influenza-specific PRO instruments also have an important role in clinical research in elucidating the efficacy and safety profiles of influenza interventions, both as primary and secondary endpoints. For example, the FLU-PRO and FluiiQ™ have been used in multiple randomized clinical trials of influenza interventions (22, 25, 29). The importance of PRO measures in determining the benefit-risk ratio of influenza interventions is highlighted by the inclusion of PRO data in the clinical section of the product labels of multiple US FDA- and EMA-approved antiviral medications (38–45). Indeed, several regulatory documents aimed at informing the design of influenza clinical trials recommend the inclusion of PROs focusing on influenza symptoms as secondary endpoints (36, 37).

There are, however, several challenges regarding use of PRO instruments in influenza research and care, including content validity and reliability, patient compliance and data collection, privacy and confidentiality, and equitable access (7, 47–50). A major challenge for successful implementation of PROs in influenza research is the selection of an appropriate instrument; it is vital to consider both the content validity (i.e. degree of how much the measure of interest is captured) and the reliability (i.e. degree of reproducibility) in the intended context of use (7), avoiding applying instrument psychometric properties out of the clinical trial setting in which they were originally assessed (15, 49). Sensitivity to change or responsiveness and thresholds for meaningful change are also important measurement characteristics to be considered (7, 49); these are particularly useful in clinical care settings where treatment decisions would be based on changes in the PRO scores (7, 15).

Factors that impact biases and confounding include the inherently subjective nature of PROs that rely on patients’ self-assessment and their interpretation of their symptoms and the impact of disease (47). Other potential biases and confounders that can make data collection and interpretation difficult include social desirability bias (where patients provide answers they deem to be socially desirable or expected), and selection bias (where patients who experience more severe symptoms or are dissatisfied with their healthcare are more inclined to participate) (49, 51). A patient’s cognitive abilities can also substantially influence their understanding of PRO questions and their ability to provide accurate responses (49). Various methods have been employed to address these content validity concerns, including ensuring cognitive interviews are used to assess understanding (48, 49), and having active involvement of participants in the development and refinement of PROs (28, 49, 50).

Patient compliance and data collection can introduce bias when using PROs. Factors that can impact data collection include non-responses and high patient dropout rates, which can impact the completeness of the data and lead to a selection bias, potentially affecting the validity of the study findings (49). Non-responses may be impacted by data collection burden, as long or frequent assessments can be burdensome for patients, leading to survey fatigue and reduced compliance (49, 50). Therefore, researchers must balance the need to collect comprehensive data with ensuring patient comfort and well-being, potentially by considering the optimal length, format, and timing of assessments to minimize the burden on participants (50). For respondents, there can be technological barriers given the increasing reliance on digital platforms for data collection; this can lead to underrepresentation of certain populations who lack access to technology or have limited digital literacy (50). Human error can also be introduced in data entry when transferring data into research databases (47). Therefore, researchers must also consider technological aspects of collection and data entry in the development and application of PRO instruments.

Given the nature of PROs, there are several ethical considerations regarding privacy and access. Privacy and confidentiality should be considered key pillars in the ethical acquisition, storage, and utilization of such data. Ensuring the protection of data and the safeguarding of patient privacy are paramount during the data collection process (47, 50). To support the privacy and confidentiality of PRO data, several measures are available to secure and protect data, including data anonymization and encryption, access restrictions, clear informed consent, and auditing and monitoring (47, 49, 50). Importantly, respecting respondent autonomy not only upholds ethical standards but also fosters trust and collaboration between researchers and participants (50). Another key ethical consideration is equitable access to PROs, i.e. ensuring that all respondents can effectively articulate their health experiences and needs (50). It is important to develop PRO measures that are culturally sensitive and relevant to a wide spectrum of patients, including ethnic, cultural, and linguistic groups (50, 52). Part of these efforts include the use of multiple languages, with contextually appropriate translations and cultural adaptations, and access to interpretation services if necessary (52, 53). Physical accessibility is also important for the equal implementation of PRO tools; for example, the provision of facilities and technologies that facilitate the participation of people with visual, hearing, or mobility impairments (50). Finally, flexible delivery of PRO tools, i.e. using paper, online, or mobile applications, empowers respondents to select the method that best aligns with their comfort and accessibility (50).

In specific populations, such as young children, the frail, or those with a disability, implementing PROs can be challenging. As such, observer-reported outcomes offer an alternative, patient-centric measure for parents and caregivers. Observer-reported outcomes provide an indirect, third-party view of the patient’s condition and potential treatment improvements, while also allowing for the measurement of caregiver burden and its impact on their quality of life (54).

Based on the findings of the systematic literature review and the authors’ clinical experience, we propose several recommendations to facilitate the wider implementation of PROs in influenza clinical research and care (Table 3).

**Table 3.** Author recommendations based on findings of the systematic review and the authors’ clinical experience.

| Recommendation | Notes |
| --- | --- |
| *Recommendation #1: Further research is needed to determine the value of healthcare providers incorporating routine PRO assessments in influenza clinics, ensuring that the patient's perspective is considered in clinical decision-making, to allow more personalized and effective care.* | Regular and routine collection of PRO data relating to patients' disease-related and vaccine-specific reactogenic symptoms, well-being, and response to treatment complements traditional influenza metrics of infection, severe disease or mortality. This could allow healthcare providers and decision-makers to personalize prophylactic and symptomatic interventions and discern between alternative strategies (6). However, further research is needed to assess the application, value and uptake of these identified influenza-specific PRO instruments in real-world influenza care scenarios. |
| *Recommendation #2: Public immunization campaigns may benefit from the use of PROs to understand and communicate the broad benefits of influenza vaccines, with the potential to increase vaccination awareness and uptake, and to implement strategies that reduce the burden of disease.* | PRO data can highlight the broad protective effects of influenza vaccines, helping to frame influenza vaccination as beneficial beyond just reducing probability of infection. For example, PRO data could be used to communicate improvements in symptom reduction, quality of life, and reduction of side effects following vaccination (28, 29, 32). Researchers and public health bodies have also employed PRO instruments to capture vaccine recipients' experiences and perceptions. For example, in the UK, researchers investigated parental attitudes towards childhood influenza vaccines, including self-reported uptake, adverse events, and intention to vaccinate in the following influenza season. These data provide important feedback on vaccination campaigns and provide information regarding factors that may reduce vaccine uptake (56). |
| *Recommendation #3: The pharmaceutical industry should reinforce the inclusion of PRO endpoints, generic- and disease-* | The SPIRIT statement (Standard Protocol Items: Recommendations for Interventional Trials) provides evidence-based recommendations for the minimum content of a clinical trial protocol and is widely endorsed as an international standard for trial protocols. In 2018, an extension of |
| *specific ones, in the development, evaluation and monitoring of influenza interventions, to inform drug development, regulatory submissions, and post-marketing surveillance.* | the SPIRIT statement (SPIRIT-PRO) was published, providing recommendations specifically for trials involving PROs as key primary or secondary outcomes. These recommendations, when integrated into clinical trials, promote the collection of high-quality PRO data that can inform patient-centered care (47).<br>Regulatory agencies, including the US FDA and EMA, have also recognized the growing importance of PROs in the evaluation of new interventions. For example, one study analyzed regulatory data published by the EMA for authorized products between 2008 and 2012. From a total of 180 identified products, 17% included PROs as the primary evaluation criterion, and PROs were used as an endpoint for 82 products (57). In influenza, the FDA has suggested the inclusion of primary and secondary endpoints for symptomatic and prophylactic interventions, respectively, for a regular measure of symptoms in those infected (36). |
| *Recommendation #4: Future research on PROs should prioritize the use of valid and reliable instruments in the intended clinical setting.* | The increasing adoption of PROs in clinical decision-making depends on the trustworthiness of the generated outcomes. As such, the employment of valid instruments is crucial as it guarantees that the collected data accurately mirror the patient's health status, thereby providing an authentic depiction of their experiences. However, it's important to note that the documented psychometric properties of existing PRO instruments are confined to the context in which they were studied. Therefore, each new implementation necessitates prior research along with case-by-case validity and reliability testing. |

EMA: European Medicine Agency, FDA: United States Food and Drug Administration, PRO: patient-reported outcome, UK: United Kingdom

## Conclusions

In conclusion, PROs provide a patient-centered perspective that complement traditional clinical metrics for influenza. This systematic literature review identified several PRO instruments that have been used in clinical trials to assess the severity of influenza symptoms and impact of influenza, as well as the acceptability and safety of influenza interventions. We also identified the inclusion of PRO results in the clinical section of label claims for various influenza interventions and FDA guidelines that recommend the inclusion of PROs in influenza clinical trial designs, highlighting the increasing importance and acceptance of PROs. However, beyond clinical trial settings, studies are needed to better understand how PROs may enhance routine care in real-world clinical settings. The results of this systematic literature review can help to inform and guide healthcare professionals, patients, decision-makers, and clinical researchers on the advantages and challenges of PROs and recommendations for the integration of such tools in influenza clinical research and care.

## Abbreviations

CARIFS: Canadian Acute Respiratory Illness and Flu Scale
COA: Clinical Outcome Assessment
EMA: European Medicines Agency
EQ-5D: EuroQol five dimensions
FDA: Food and Drug Administration
Hemo-TEM: Hemophilia Treatment Experience Measure
FLU-PRO: InFLUenza Patient-Reported Outcome
FluiiQ™: Influenza Intensity and Impact Questionnaire
ILI: Influenza-like illness
ISS: Influenza symptom severity
IIWS: Influenza impact wellbeing scale
MeSH: Medical Subject Heading
PRO: Patient-reported outcomes
QoL: Quality of life
RiiQ™: Respiratory Infection Intensity and Impact Questionnaire
SF-36: 36-item short form health survey
VAPI: Vaccinees’ Perception of Injection

## Declarations

## Acknowledgements

Medical writing support under the guidance of the authors was provided by Phoebe Liddell of Ashfield MedComms, an Inizio company, and funded by Sanofi. Editorial support was provided by Olivia Morris of Ashfield MedComms, an Inizio company, and funded by Sanofi.

## Author contributions

EMM and PMA: Performed the systematic literature review, wrote the manuscript, and reviewed and approved changes.

BAFM, CG, JBH: Participated in the critical assessment of literature findings, wrote sections of the manuscript, and reviewed the accuracy of the information.

## Data availability

This is a review paper and thus no experimental or clinical data have been generated.

## Ethics approval

Not applicable.

## Consent for publication

Not applicable.

