## Supplementary File for "Raising the patient’s voice in influenza research and care with patient-reported outcomes"

**Supplementary File S1**

**Validity and reliability of patient-reported outcomes**

Ensuring the validity of patient-reported outcomes (PRO) is imperative to ascertain that the instrument effectively encompasses the pivotal concepts of interest it aims to gauge (1). This validation process entails a comprehensive qualitative phase involving patient engagement, which serves as the cornerstone for ensuring the measure's appropriateness within the target population affected by the disease.

The steps involved in assessing PRO validity encompass face, construct, convergent, divergent, and criterion validity. While detailed documentation can be found at the provided references, a general overview of these validation steps is provided below (1):

i. Face validity offers an initial, overarching assessment of the instrument's suitability in relation to the disease context, typically without a standardized procedure.
ii. Construct validity ensures alignment between the abstract way how individuals characterize disease-attributes (constructs) and the actual items being measured in the instrument wording.
iii. Divergent validity ensures the sensitivity of the scale when measuring distinct groups
     (e.g. based on age or severity).
iv. Criterion validity compares the scores of the instrument with those of a gold standard.

In addition to instrument validity, reliability addresses the precision level of the instrument in capturing the concept of interest devoid of measurement errors (1). Two reliability measures are commonly utilized: internal consistency reliability and test-retest reliability.

i. Internal consistency reliability evaluates the correlation between items when more than one is included.
ii. Test-retest reliability ensures consistency in results, by repeatedly assessing the same individuals’ scores at different time points.

The Systematic Literature Review identified four influenza-specific papers that describe, to varying extents, the validity and reliability of existing PRO instruments. Understanding the context underlying these studies, and the level of evidence presented, is pivotal in assessing the suitability of the tool across diverse contexts. **Table S1** provides a non-exhaustive evaluation of the psychometric properties outlined in these papers. The Influenza Symptom Severity (ISS) scale and Influenza Impact Wellbeing Scale (IIWS), both of which have been discontinued, were subsequently incorporated into the FluiiQ™ after extensive psychometric analysis and are herein not discussed.

**Table S1** General psychometric characteristics of available PRO instruments for influenza

| **Instrument and  papers assessed** | **Psychometric Considerations** | |
| --- | --- | --- |
| **Influenza Intensity and Impact Questionnaire (FluiiQ™)**  *Osborne RH, et al. 2011 (2)* | Sample size and respondent selection: | The study encompassed 159 participants with ILI and 75 with LCI across various medical care levels and multiple locations in Australia and the United States. Age range varied between 19 to 55 years old. |
|  | Psychometric properties: | The instrument demonstrated strong reliability in four out of five domains and provided evidence of criterion and divergent validity. However, further research is required to assess test-retest reliability, considering the evolving nature of influenza symptoms over time. |
|  | Generalizability: | The instrument underwent testing at diverse sites, medical care levels and diverse age populations. Furthermore, the paper discusses its utilization in multinational clinical trials. While this evidence can offer guidance for the instrument's application in various contexts, it also suggests that the validity of the instrument should be evaluated on a  case-by-case basis. |
| **InFLUenza Patient-Reported Outcome (FLU-PRO)**  *Powers JH, et al. 2016 (3), Powers JH, et al. 2018 (4)* | Sample size and respondent selection: | The initial phase involved 34 ILI adult participants aged between 37 and 39 years old from the United States and Mexico, encompassing various clinical sites. Subsequently, a second study recruited 220 ILI patients from the United States and other countries, with a comparable age distribution. |
|  | Psychometric properties: | The study exhibited strong internal and test-retest reliability across domains. Validity assessments, including construct and divergent analyses, demonstrated the instrument's ability to discern levels of severity in different populations. |
|  | Generalizability: | The evidence presented offers valuable guidance for conducting similar research in populations akin to those enrolled in the trials, not discarding the importance of reconducting study-specific validation steps. However, the narrow age range of patients may constrain the extrapolation of findings, especially when examining both younger and older adult populations. |
| **Vaccinees' Perception of Injection (VAPI)**  *Chevat C, et al. 2009 (5)* | Sample size and selection: | During the initial development phase, 549 patients aged 18 to 74 years were involved, while the psychometric validation phase included  5543 healthy adult subjects from various European countries with a mean age of 57 years. |
|  | Psychometric properties: | VAPI demonstrated robust internal reliability across all age groups and underwent psychometric validation, encompassing assessments of construct and face validity. It furthermore claims evidence of its use in the clinical practice. However, limitations exist, notably the absence of test-retest reliability and the lack of confirmatory factor analysis prior to utilization. This omission stems from potential biases in item fitting to scales attributable to sentence structure. |
|  | Generalizability: | The generalizability of study findings may be constrained by the limited information available on elderly patients aged 57 and above, as well as the geographic coverage. Presented estimates regarding the perception of conventional intramuscular vaccination among similar individuals can establish a baseline for future studies. |

ILI: influenza-like illness, LCI: laboratory-confirmed infection
